# The founder missense mutation of *WFDC2* in Koreans leads to severe respiratory distress accompanied by bronchiectasis and rhinosinusitis

**DOI:** 10.1101/2024.11.16.24317083

**Authors:** Jae Won Roh, Jiyoung Oh, Se Jin Kim, Ji Won Hong, Jiyeon Ohk, Hyo Sup Shim, Hyung-Ju Cho, Ala Woo, Song Yee Kim, Young Ae Kang, Hosung Jung, Kyung Won Kim, Moo Suk Park, Heon Yung Gee

**Affiliations:** Department of Pharmacology, Graduate School of Medical Science, Brain Korea 21 Project, Yonsei University College of Medicine, Seoul 03722, Republic of Korea; Woo Choo Lee Institute for Precision Drug Development, Seoul 03722, Republic of Korea; Department of Pediatrics, Severance Hospital, Yonsei University College of Medicine, Seoul 03722, Republic of Korea; Department of Anatomy, Graduate School of Medical Science, Brain Korea 21 Project, Yonsei University College of Medicine, Seoul 03722, Republic of Korea; Department of Pathology, Yonsei University College of Medicine, Seoul, Korea; Department of Otorhinolaryngology, The Airway Mucus Institute, Korea Mouse Phenotyping Center (KMPC), Taste Research Center, Yonsei University College of Medicine, Seoul 03722, Republic of Korea; Department of Internal Medicine, Severance Hospital, Yonsei University College of Medicine, Seoul 03722, Republic of Korea

**Author notes:** Correspondence: Kyung Won Kim, Department of Pediatrics, Yonsei University College of Medicine, 50-1 Yonsei-Ro, Seodaemun-gu, Seoul 03722, Korea, Moo Suk Park, Department of Internal Medicine, Yonsei University College of Medicine, 50-1 Yonsei-Ro, Seodaemun-gu, Seoul 03722, Korea, Heon Yung Gee, Department of Pharmacology, Yonsei University College of Medicine, 50-1 Yonsei-Ro, Seodaemun-gu, Seoul 03722, Korea. These authors contributed equally (Jae Won Roh, Jiyoung Oh, Se Jin Kim, Ji Won Hong).

## Abstract

**Background:** Chronic airway diseases like cystic fibrosis (CF) and primary ciliary dyskinesia (PCD) pose substantial clinical challenges. This study explores the p.C97W variant in *WFDC2*, proposed as a new genetic origin of respiratory distress, especially among Koreans.

**Methods:** Whole-exome/genome sequencing (WES/WGS) were performed on 64 patients from 62 families presenting with severe bronchiectasis and chronic rhinosinusitis. *In vitro* analyses and protein modeling were utilized to evaluate the functional implications of the *WFDC2* variant.

**Results:** Pathogenic variants were found in 11.3% of families, including a novel homozygous *WFDC2* missense variant (c.291C>G, p.Cys97Trp) in five unrelated families, confirmed by Sanger sequencing. The variant, rare globally but more frequent in Koreans, caused persistent wet cough, chronic rhinosinusitis, and bronchiectasis in affected patients. Lung tissue pathology showed chronic inflammation and interstitial fibrosis. The p.C97W variant impaired WFDC2 protein folding, secretion, and function.

**Conclusions:** The p.C97W variant in *WFDC2* is a critical genetic factor in severe chronic airway disease that shares clinical features with CF and PCD. Given its implications for diagnosis and treatment, genetic testing for *WFDC2* mutations in individuals with CF or PCD-like symptoms is recommended.

## INTRODUCTION

In humans, the respiratory epithelium is critical for air transport, mucociliary clearance, and immune regulation. The airway epithelium comprises various cell types, including basal, ciliated, goblet, and club cells, as well as rare types like tuft cells, ionocytes, and neuroendocrine cells.^1^ The mucosal surface of the respiratory tract contributes to innate immune defense. Each epithelial cell type plays a distinct role in maintaining airway integrity and the mucociliary clearance system, by producing antioxidants, protease inhibitors, antimicrobial peptides, or by secreting mucins and surfactants.^2,3^ Dysregulation of these processes due to genetic or environmental factors can lead to the pathophysiology of severe viral infections and chronic pulmonary disorders, including asthma,^1,4,5^ cystic fibrosis (CF),^1,6-8^ and primary ciliary dyskinesia (PCD).^1,9,10^

In this study, we identified a homozygous missense variant (c.291C>G, p.C97W) in *WFDC2*, a serine/threonine protease inhibitor with anti-bacterial activity, through whole-exome and genome sequencing in families affected by respiratory conditions resembling PCD. Individuals with this variant exhibited chronic inflammation, bronchiectasis, interstitial fibrosis, and infections caused by *Pseudomonas aeruginosa*. Due to its implications for diagnosis and treatment, genetic testing for WFDC2 mutations is recommended for individuals with CF or PCD-like symptoms.

## RESULTS

### Identification of the *WFDC2* variant

The WES or WGS analysis pipeline to identify the potential genetic causes in patients with severe bronchiectasis and chronic rhinosinusitis (n = 64 from 62 families) is shown in Fig. S1. Pathogenic variants were identified in 7 of the 62 families (11.3%): five with variants in PCD-linked genes (six patients) and two (two patients) with CFTR variants (Fig. 1A, B). Furthermore, a homozygous missense variant (c.291C>G, p.Cys97Trp) in *WFDC2* was identified in five unrelated families (Fig. 1C, and Fig. S2). Sanger sequencing confirmed this homozygous mutation in affected probands and their siblings, while the unaffected mother was heterozygous (Fig. 1D). The allele frequency of this variant was 0.00001611 in the global population 0.0004051 in East Asians, and 0.0018 in the Korean population, as reported in the Genome Aggregation Database (gnomAD; Table S1). Haplotype analysis revealed that individuals carrying the c.291C>G *WFDC2* mutation shared a genomic region of approximately 138.5 kb surrounding the variant (Table S2). Subsequent haplotype analysis using the LDhap Tool^11^ using eight selected SNPs within the shared region revealed that individuals with the c.291C>G *WFDC2* mutation share a rare H4 haplotype, with a frequency of 0.63% in the homozygous state (Table S2). This finding suggests this variant as a founder allele.

**Fig. 1.**
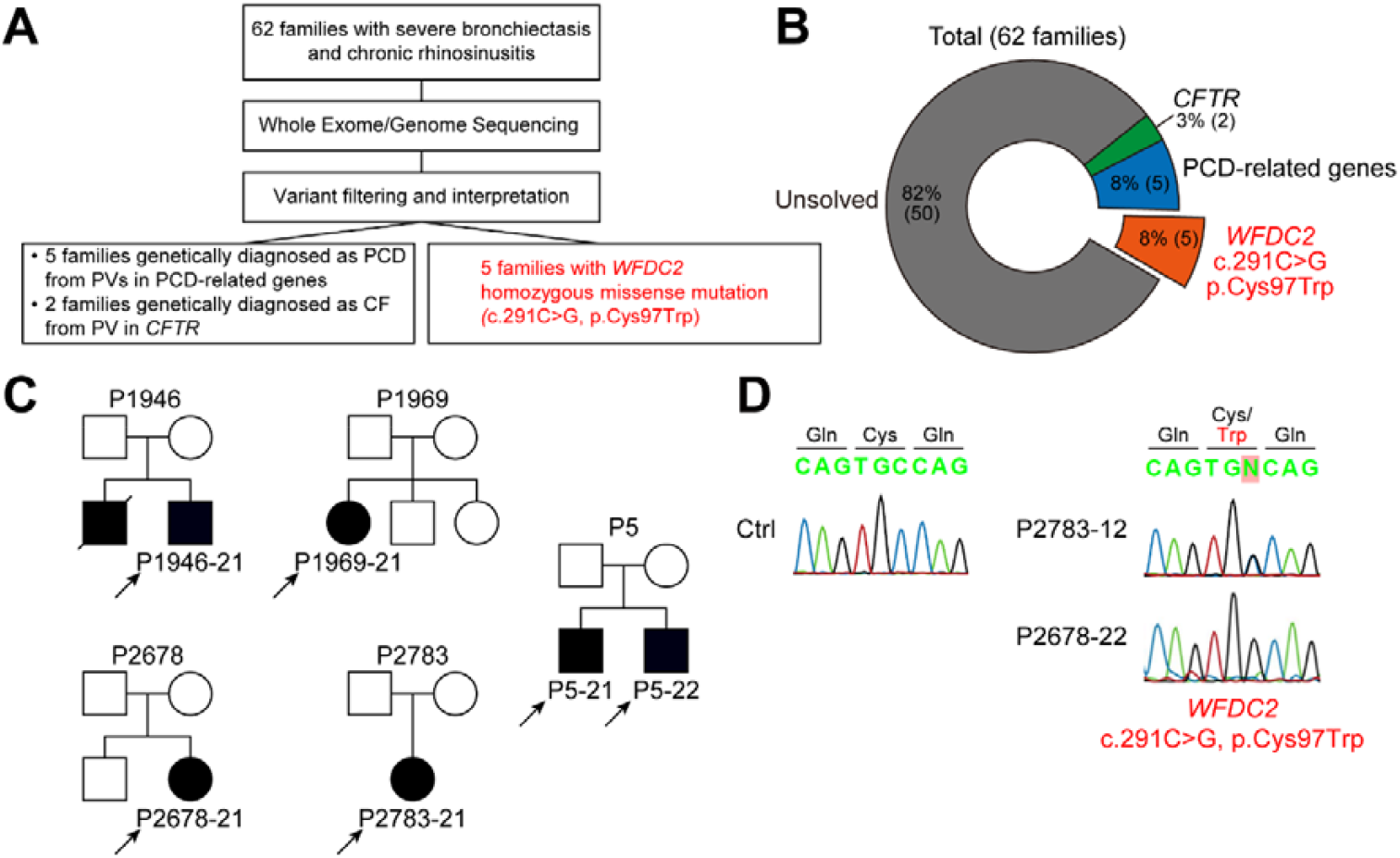
Identification of a homozygous variant in *WFDC2* in families with severe bronchiexctasis and chronic rhinosinusitis. (A, B) Summary and result of WES/WGS analysis to elucidate the causal variants in patients with severe bronchiectasis and chronic rhinosinusitis. Five families had a homozygous variant in *WFDC2*, which was the c.291C>G substitution resulting in the amino acid change p.Cys97Trp. (C) Pedigree of five families with the c.291C>G *WFDC2* variant. The probands are indicated using arrows. (D) Sanger sequencing result of c.291C>G *WFDC2* variant representing individuals from three families. The altered amino acid is written in red. WES, Whole exome sequencing; WGS, whole genome sequencing.

### Clinical phenotypes of individuals with the *WFDC2* variant

All six patients from five unrelated families presented with persistent wet cough and chronic rhinosinusitis (Fig. 2C). Chest CT scans revealed bronchiectasis in five patients, while one patient (P1946-21) declined the scan (Fig. 2A). Pathological examination of the original lung tissue from three female patients (P1969-21, P2678-21, and P2783-21) who underwent lung transplantation at an average age of 33 years (range: 21–41 years; Table 1) revealed chronic inflammation, bronchiectasis, and interstitial fibrosis (Fig. 2D). All patients had *P. aeruginosa* lung infections, and pulmonary function demonstrated a severe airflow obstruction pattern before lung transplantation. Two siblings (P5-21 and P5-22) of three male patients recently visited our hospital, both presenting with bronchiectasis and reduced FEV1% predicted (Fig. 2B, and Table 1).

**Table 1.** Clinical characteristics of the patients.

| Patient | P1946-21 | P1969-21 | P2678-21 | P2783-21 | P5-21 | P5-22 |
| --- | --- | --- | --- | --- | --- | --- |
| Age at confirmation of *WFDC2* mutation, years | 31-35 | 36-40 | 46-50 | 31-35 | 21-25 | 21-25 |
| Sex | Male | Female | Female | Female | Male | Male |
| Bronchiectasis | N/A | Yes | Yes | Yes | Yes | Yes |
| *Psuedomonas aeruginosa* infection | N/A | Yes | Yes | Yes | N/A | N/A |
| Chronic rhinosinusitis | Yes | Yes | Yes | Yes | Yes | Yes |
| Nasal polyposis | No | No | No | Yes | Yes | Yes |
| Age at transferred, years | 26-30 | 31-35 | 36-40 | 16-20 | 21-25 | 21-25 |
| Lung Transplantation (age, years) | No | Yes (36-40) | Yes (41-45) | Yes (21-25) | No | No |
| Medical history of siblings | Deceased due to pneumonia | No | Reduced lung function | No | Sibling of P5-22 | Sibling of P5-21 |
| Pulmonary function * |  |  |  |  |  |  |
| FEV<sub>1</sub> (% pred) | N/A | 29 | 21 | 22 | 85 | 71 |
| FEV<sub>1</sub> (L) | N/A | 0.86 | 0.56 | 0.69 | 3.55 | 3.14 |
| FEV<sub>1</sub>/FVC (%) | N/A | 44 | 49 | 44 | 81 | 81 |
| DLCO |  | 37 |  |  |  |  |

N/A, data not available
\* Pulmonary function tests were performed just before lung transplantation for P1969-21, P2678-21, and P2783-21, and most recently for P5-1 and P5-2.

**Fig. 2.**
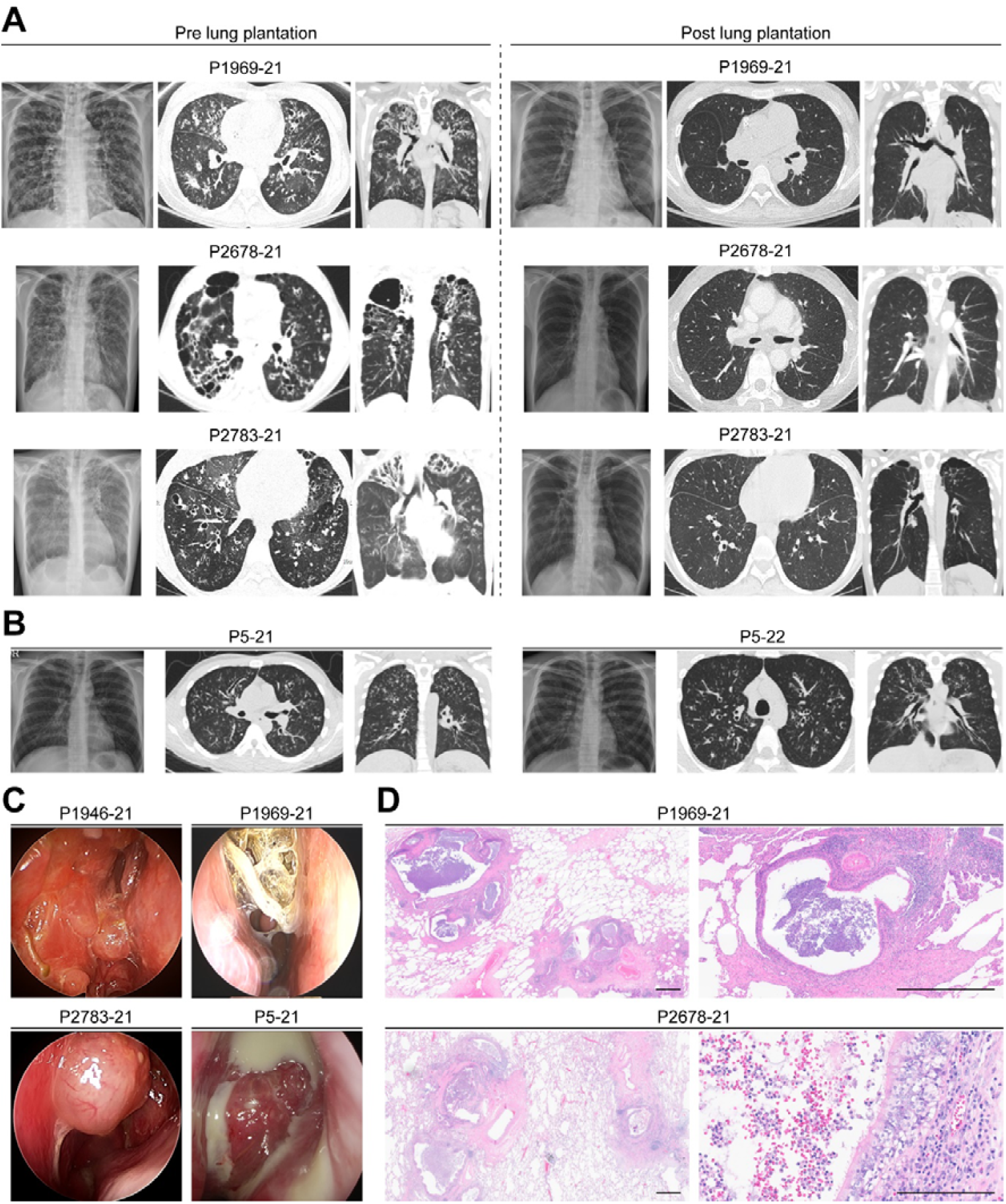
Clinical manifestation of individuals with bi-allelic *WFDC2* variant. (A) Overall structure of the WFDC2 protein. Two WAP domains and the disulfide bond-forming cysteines are shown. C97 forms a disulfide bond with C109. (B) Multiple sequence alignment of human WFDC2 against its orthologs from five different vertebrate species was performed using Jalview.^19^ The mutated amino acid residue (C97) and the disulfide bond-forming cysteine residue with C97 (C109) are indicated. (C) Monomer and homo-dimer structure prediction of WFDC2 WT and p.C97W mutant by AlphaFold2.^12,13^ (D) Western blot analysis of WT and p.C97W mutant WFDC2 proteins in both the lysates and secreted media of HEK293T cells overexpressing the proteins. In (D), data are presented in mean ± standard error of the mean. In (D), a two-way Student’s t-test was used to calculate the p-value, and a p-value < 0.05 was considered significant. *, p < 0.05; ****, p < 0.0001.

The average age at the onset of respiratory symptoms was 7.8 years (range, 1 month to 13 years). Initial clinical diagnoses based on respiratory symptoms included neonatal pneumonia in patients P1946-21 and P1969-21; bronchiectasis in patients P2678-21, P2783-21, P5-21, and P5-22; and chronic rhinosinusitis in patients P5-21 and P5-22. Bronchiectasis was confirmed during childhood in two-thirds of the patients, with one patient diagnosed at late 0’s (P2678-21) and three patients diagnosed at early 10’s (P2783-21, P5-21, and P5-22). The three patients who underwent lung transplants (P1969-21, P2678-21, and P2783-21) have maintained stable lung structure and function for two, seven, and fourteen years, respectively (Fig. 2, and Table 1).

### Effect of p.C97W variant on WFDC2 function

As shown in Fig. 3A, WFDC2 contains a N-terminal signal peptide that is involved in trafficking and secretion, and two whey acidic protein (WAP) domains connected by a short loop. Each WAP domain includes eight cysteine residues that form four disulfide bonds. Structure prediction using AlphaFold2^12^ showed that C97 located in the second WAP domain forms a disulfide bond with C109 (Fig. 3A). Multiple sequence alignments of human WFDC2 with its orthologs from several vertebrate species demonstrated that both C97 and C109 are highly conserved across vertebrates (Fig. 3B). Additionally, the *in silico* prediction scores classified the C97W variant as pathogenic or disease-causing (Table S1).

**Fig. 3.**
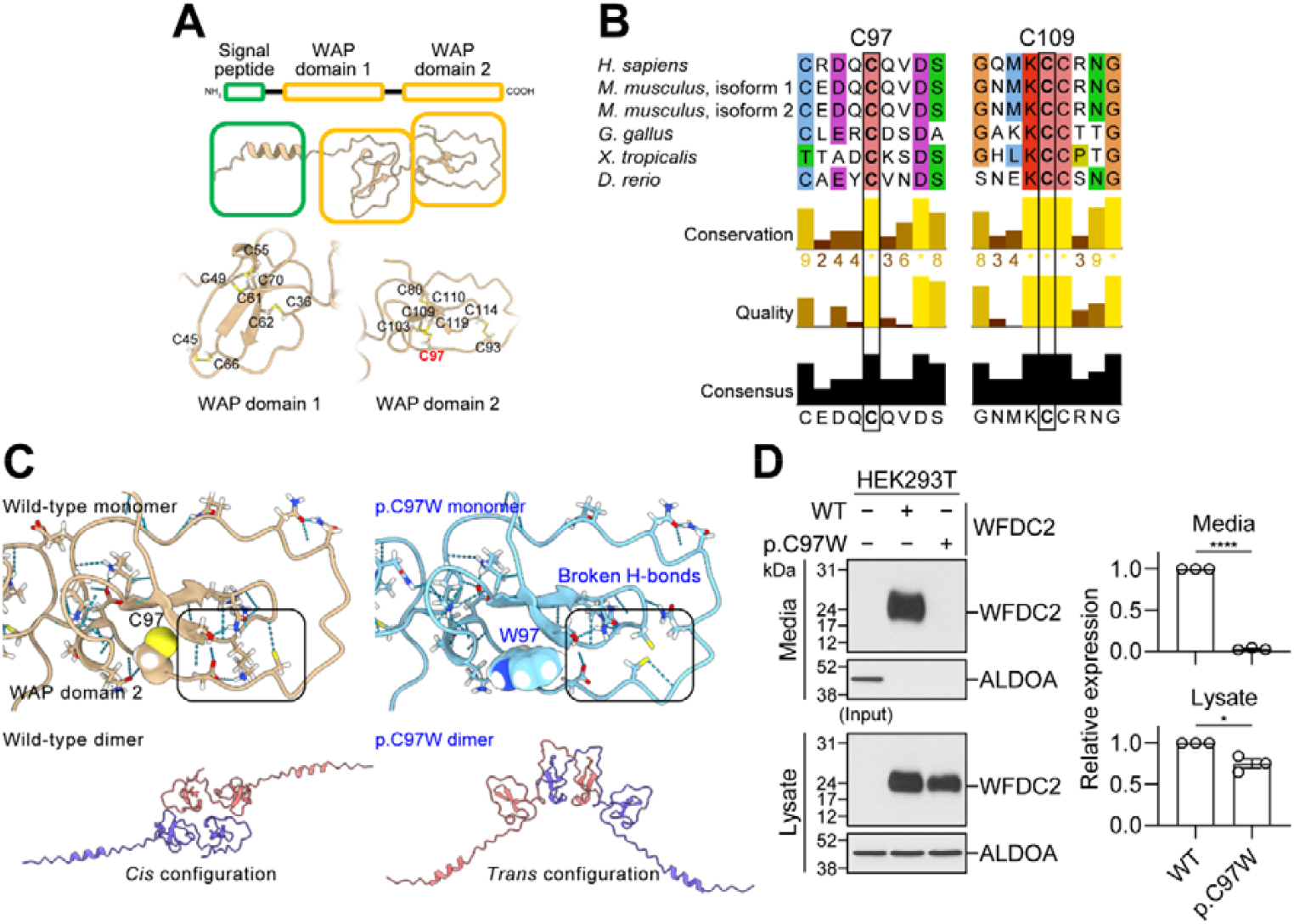
Molecular mechanism of *WFDC2* p.C97W mutant. (A) Overall structure of the WFDC2 protein. Two WAP domains and the disulfide bond-forming cysteines are shown. C97 forms a disulfide bond with C109. (B) Multiple sequence alignment of human WFDC2 against its orthologs from five different vertebrate species was performed using Jalview.^19^ The mutated amino acid residue (C97) and the disulfide bond-forming cysteine residue with C97 (C109) are indicated. (C) Monomer and homo-dimer structure prediction of WFDC2 WT and p.C97W mutant by AlphaFold2.^12,13^ (D) Western blot analysis of WT and p.C97W mutant WFDC2 proteins in both the lysates and secreted media of HEK293T cells overexpressing the proteins. In (D), data are presented in mean ± standard error of the mean. In (D), a two-way Student’s t-test was used to calculate the p-value, and a p-value < 0.05 was considered significant. *, p < 0.05; ****, p < 0.0001.

Next, we predicted the structures of both the wild-type (WT) and p.C97W mutant *WFDC2* using ColabFold^13^ and AlphaFold2.^12^ The p.C97W mutant failed to form a disulfide bond with C109, in contrast to WT (Fig. 3C). Additionally, the bulky side chain of the Trp (W) residue disrupted hydrogen bond formation near W97 in the second WAP domain (Fig.3C). WFDC2 forms dimeric structures;^14^ therefore, we predicted the dimeric structures of WT and p.C97W. Clustering of the structures from multiple AlphaFold2 runs showed that the p.C97W structure predominantly exists in a ‘trans–*trans*’ configuration, where the second WAP domain is untangled and interacts with the other subunit. In contrast, the WT WFDC2 dimers were more likely to exist in the ‘*cis*’ configuration (Fig. 3C, and Fig. S3).

Assessment of the effects of the p.C97W mutant on WFDC2 secretion in human embryonic kidney (HEK293T) cells overexpressing WT or p.C97W WFDC2 revealed that WT WFDC2 produced a single band of approximately 24 kDa in both the lysate and culture media. In contrast, the p.C97W variant was not secreted and showed reduced expression in the lysate (Fig. 3D). Immunofluorescence analysis in HEK293T cells showed colocalization of both WT and p.C97W WFDC2 with the endoplasmic reticulum marker, suggesting no defect in intracellular localization (Fig. S4A). Furthermore, treatment with tunicamycin, an inhibitor of N-linked glycosylation, or glycosidase digestion by incubation of cell lysates with glycosidases, such as PNGase F or Endo H, demonstrated that both WT and p.C97W were glycosylated (Fig. S4B, C), as indicated by the shift in band molecular weight with no difference between the WT and mutant (Fig. S4B, C). This finding indicated that WFDC2 was glycosylated, as previously described.^14^ In summary, these findings indicated that the p.C97W mutation disrupts WFDC2 protein folding and secretion, impairing its function.

## DISCUSSION

In this study, we identified a novel *WFDC2* mutation (c.291C>G; p.C97W) as the causative factor in severe lung disease. This autosomal recessive variant leads to chronic inflammation, bronchiectasis, interstitial fibrosis, and severe airflow obstruction, often necessitating lung transplantation. Population data from gnomAD and KRGDB indicate that this allele has a relatively high frequency in East Asian populations, particularly in the Korean population. Based on these data, approximately three out of one million individuals are homozygous for this mutation, suggesting that around 150 individuals in South Korea could be affected. In contrast, the previously reported c.145T>C; p.C49R variant, identified as a European founder allele, is extremely rare in East Asian populations, including Koreans. However, it has a higher frequency in the European population with an allele frequency of 0.0001814, equivalent to three individuals per ten million and approximately 2,239 patients in Europe. Additionally, 22 loss-of-function (LoF) alleles in *WFDC2* have been reported in gnomAD, with a combined homozygous frequency of approximately 0.0000000003178.

Respiratory phenotypes of WFDC2 deficiency identified in our study and another study^15^ present with clinical features that closely resemble CF and PCD, with symptoms including bronchiectasis, chronic rhinosinusitis, and recurrent respiratory infections. Like CF, patients with *WFDC2* mutations develop bronchiectasis across all lung lobes and are prone to chronic *P. aeruginosa* infections, indicating a more severe and diffuse impact on lung structure in WFDC2 deficiency and CF compared to PCD. Furthermore, nasal and sinus abnormalities, including chronic rhinosinusitis and nasal polyps, are prevalent across all three conditions. In this study, three patients with *WFDC2* mutations underwent lung transplantation due to respiratory failure, further highlighting the severe progression of the disease.

The *WFDC2* c.291C>G; p.C97W variant shares molecular similarities with the previously described c.145T>C; p.C49R variant.^15^ Both are autosomal recessive missense mutations in cysteine residues within the WAP domain, a critical region for disulfide bond formation, and lead to LoF by disrupting the WAP domain. The key difference between the two variants is that the p.C49R variant affects the first WAP domain, while the p.C97W variant affects the second. Additionally, unlike the p.C49R variant, which is associated with glycosylation defects,^15^ our data did not reveal glycosylation differences for the p.C97W variant, nor did it alter intracellular localization compared to the WT protein. Structural modeling using AlphaFold2,^12^ aligns well with the disruption of WAP domain integrity due to the loss of disulfide bonds, suggesting that improper dimer formation might explain the differences in the effects of the p.C97W variant. Despite these structural differences, both variants exhibit similar defects in protein secretion, suggesting comparable pathological consequences in humans.

In conclusion, we identified the p.C97W *WFDC2* variant as a novel cause of severe chronic airway disease, which is physiologically distinct but clinically similar to CF and PCD. Our study revealed the prevalence of the *WFDC2* p.C97W variant in the Korean population, suggesting genetic testing for this gene could be beneficial in patients who have respiratory phenotypes resembling PCD or CF.

## METHODS

### Study design

This study was approved by the Institutional Review Board of Severance Hospital, Yonsei University Health System (IRB #4-2013-0770 and #4-2024-0647). Written informed consent was obtained from the patients for their participation in this study and the publication of their clinical data. Sixty-four patients with severe bronchiectasis and chronic rhinosinusitis from 62 unrelated families were enrolled from both the PCD-like severe bronchiectasis and chronic rhinosinusitis and lung transplantation cohorts at a single tertiary medical center in South Korea. These patients met the European Respiratory Society guidelines for the clinical diagnosis of PCD,^16^ which include symptoms such as persistent wet cough, chronic rhinosinusitis, or bronchiectasis. Whole-exome sequencing (WES) or whole-genome sequencing (WGS) was performed to identify potential genetic causes. In-depth genetic analyses were performed on individuals without any potential molecular causes.

### Analysis of whole exome/genome sequencing data

Genomic DNA was extracted from blood samples of patients clinically suspected of having PCD using a DNeasy Kit (Qiagen, Germany). Whole exome sequencing (WES) was performed using a SureSelect V5 kit (Agilent Technologies, Santa Clara, CA, USA) and an Illumina HiSeq 2500. Raw WES FASTQ files were imported into CLC Genomics Workbench version 9.5.3 for data analysis. Sequenced reads were mapped to the human reference genome, GRCh37/hg19. Variants were detected using the Basic Variant Caller of the CLC Genomics Workbench, with a minimum coverage of 5, a count of 2, and a frequency of 20%. Low-coverage whole genome sequencing (WGS) was performed using a PCR-free DNBSEQ platform (BGI, Beijing, China). Following GATK best practice guidelines, the reads were aligned to the human reference genome GRCh37/hg19 using the Burrows-Wheeler Aligner (BWA-MEM). Duplicates were marked, reads were sorted, and the mapping quality was recalibrated using PICARD-MarkDuplicates and GATK-BQSR. Variant calling of single nucleotide variants (SNVs) and small insertions and deletions (indels) was performed using GATK-HaplotypeCaller, and low-quality variants with a depth of coverage (DP) < 5 and genotype quality (GQ) < 20 were filtered out. Copy Number Variation (CNV) analysis was performed using EXCAVATOR version 2.2 and ExomeDepth (version 1.1.10) with default settings for the WES data. For WGS data, CNVnator (v0.3.2,https://github.com/abyzovlab/CNVnator) and GATK-SV (https://github.com/broadinstitute/gatk-sv) were used to detect CNV. Mitochondrial variants and transposable element insertion calling were performed using Mutect2 and xTea (v0.1.9; https://github.com/parklab/xTea).

SNVs and small indels were annotated using Variant Annotation tools in the CLC Workbench or ANNOVAR software. SNVs and small indels were first filtered based on minor allele frequency (MAF), applying a threshold of MAF < 1% according to the Single-Nucleotide Polymorphisms Database (dbSNP), Genome Aggregation Database (gnomAD), and Korean Reference Genome Database (KRGDB); variants that were nonsynonymous or located in splice sites were selected. To evaluate pathogenicity and prioritize the candidate causing variants, *in silico* prediction algorithms, such as MutationTaster, Sorting Tolerant from Intolerant (SIFT), Combined Annotation Dependent Depletion (CADD), Polymorphism Phenotyping v2 (PP2) scores, and public databases including Online Mendelian Inheritance in Man (OMIM) and ClinVar, were used. Interpretation of the variants was performed according to the American College of Medical Genetics and Genomics (ACMG) guidelines.

To identify splice site variants, SpliceAI was run on the variants using Ensembl’s Variant Effect Predictor (VEP) tool and filtered using a threshold MAF of 1% and delta score threshold of 0.1. Variants were annotated based on their presence in human candidate cis-regulatory elements, as identified using the SCREEN database from the Encyclopedia of DNA Elements (ENCODE) project. Confirmation of the detected variants was performed manually using the Integrative Genomics Viewer (IGV) and through segregation analysis using Sanger sequencing.

### Structural prediction and clustering

AlphaFold2^12^ and ColabFold^13^ were used to construct the *de novo* monomer and dimer WFDC2 structures, respectively. The AlphaFold2-multimer-V2 algorithm was applied without using a template or minimization, using three recycle number parameters in ColabFold. The full-length WT or p.C97W human WFDC2 sequence was used as input. To generate different structural conformations, multiple runs were conducted using different seed values in ColabFold software. A total of 100 different structures were generated for the WT monomer. C97W, homodimer WT, and p. C97W. The monomer and dimer structures were then structurally aligned to perform principal component analysis (PCA) using the bio3d package^17^ in R. The *K*-means algorithm was implemented to cluster the structures. The optimal number of clusters suggested by R. ChimeraX^18^ was used to visualize protein structures.

### Cell culture, plasmids, and transfection

Human embryonic kidney (HEK) 293T cells (#CRL-3216, American Type Culture Collection, Manassas, VA, USA) were cultured in Dulbecco’s modified Eagle’s medium (DMEM) supplemented with 10% fetal bovine serum and penicillin (50 IU/mL)/streptomycin (50 μg/mL; Invitrogen, Waltham, MA, USA). The cells were incubated in a 5% CO_2_ environment at 37°C.

Plasmids encoding human *WFDC2* (NM_006103.4) and mouse *Wfdc2* (NM_026323.2, isoform 1 and NM_001374655.1, isoform 2) were purchased from OriGene (Rockville, MD, USA). The coding sequences were amplified by PCR and subcloned into the pcDNA3.4 vector using XbaI and AgeI sites. During PCR, 10xHIS tags were inserted directly into the 3′ end in front of the stop codon. Human and mouse *WFDC2* mutants were introduced using the QuikChange mutagenesis method (Agilent Technologies). The pEYFP-ER plasmid was purchased from NovoPro Biosciences (San Diego, CA, USA).

HEK293T cells were seeded in 6-well plates the day before transfection. Plasmids were transfected using a PEI-MAX transfection reagent (Polysciences, Warrington, PA, USA) according to the manufacturer’s protocol. A total of 0.5 μg of the desired plasmid and 2 μL of PEI-MAX was used for transfection. Cells were used for experiments 24–48 h post-transfection.

### Immunoblotting

Forty-eight hours after transfection, HEK293T cells were washed twice with phosphate-buffered saline (PBS) and harvested in a lysis buffer containing 150 mM NaCl, 50 mM Tris-HCl (pH 7.4), 1 mM EDTA, 1% Triton X-100, and 1x protease inhibitor cocktail (#04693116001, Sigma-Aldrich, St. Louis, MA, USA). The cells were then briefly sonicated to break the cell membrane and homogenize the sample. Bradford assay was performed to measure the protein concentration at an absorbance of 595 nm. Cell lysates (40–60 μg) were used for analysis.

The samples were incubated in 2x sodium dodecyl sulfate (SDS) sample buffer containing dithiothreitol (DTT, 0.02 g/mL) for 30 min at room temperature and subsequently separated using SDS-polyacrylamide gel electrophoresis (PAGE). The separated proteins were transferred onto a nitrocellulose membrane (GE Healthcare, Chicago, IL, USA), which was blotted using suitable primary antibodies and horseradish peroxidase-conjugated secondary antibodies in 5% skimmed milk. Protein bands were visualized using the SuperSignal West Pico kit (Thermo Fisher Scientific) and a blue film (Afga, Belgium). The density of each protein band was quantified using the ImageJ software (https://imagej.net/ij/).

To detect secreted WFDC2 proteins, transfected cells were incubated in a serum-free medium for 24 h. Culture media were collected and concentrated using Amicon Ultra-4 filters (Millipore, Burlington, MA, USA). The concentrated media was directly heated with 2x SDS sample buffer with DTT and subjected to SDS-PAGE.

The following primary and secondary antibodies were used: anti-HIS (#2365, Cell Signaling Technologies, Danvers, MA, USA; 1:1000 dilution), anti-ALDOA (#sc-390733, Santa Cruz Biotechnology, Santa Cruz, TX, USA; 1:000 dilution), HRP Anti-beta Actin antibody (#ab20272, Abcam, Cambridge, UK; 1:2000 dilution), goat anti-rabbit IgG polyclonal antibody (HRP conjugate) (#ADI-SAB-300-J, Farmingdale, Enzo Life Sciences, NY, USA), and goat anti-mouse IgG F(ab’)2 polyclonal antibody (HRP conjugate) (#ADI-SAB-100-J; Enzo Life Sciences; 1:1000 dilution).

### Immunocytochemistry

Transfected HEK293T cells were cultured on an 18 mm round coverslip. Cells were washed twice with PBS, fixed with 4% paraformaldehyde for 10 min, again washed twice with PBS, and permeabilized with 0.15% Triton X-100 in PBS for 10 min. The cells were washed twice with PBS and incubated with a blocking buffer consisting of 5% donkey serum for 1 h at room temperature. After blocking, the cells were incubated with the appropriate primary antibodies for 1 h at room temperature and washed three times with PBS. Subsequently, the cells were stained with secondary antibodies conjugated to a fluorescent marker for 1 h at room temperature and washed thrice with PBS. The samples were mounted onto glass slides using a mounting medium (Agilent Dako, CA, USA), and images were captured using a confocal microscope (LSM 700, Carl Zeiss, Germany) equipped with a 40× objective lens.

The following primary and secondary antibodies were used: anti-HIS (#2365, Cell Signaling Technologies; 1:200 dilution), Alexa Fluor 594 donkey anti-rabbit (#A21207, Invitrogen; 1:400 dilution), Alexa Fluor 647 Phalloidin (#A22287, Invitrogen; 1:400 dilution).

### Deglycosylation assay

Two methods were used to determine the glycosylation state of WFDC2. After transfection, cells were treated with 2 μM tunicamycin (Sigma-Aldrich) for 20 h. A higher dose or longer incubation time with tunicamycin caused excessive cell death, preventing further analysis. The cells were harvested and subjected to SDS-PAGE, followed by immunoblotting. Transfected cells were denatured in 2x SDS sample buffer containing DTT. Denatured samples were incubated with PNGase F or Endo H (New England Biolabs, Ipswich, MA, USA) according to the manufacturer’s protocol. Deglycosylated samples were analyzed using SDS-PAGE and immunoblotting. The shift in protein size was analyzed using both methods.

## Data Availability

All data produced in the present work are contained in the manuscript.

## ACKNOWLEDGEMENTS

We thank the families that participated in this study. We also thank the Yonsei Advanced Imaging Center and Carl Zeiss Microscope.

