## Supplementary material for "The founder missense mutation of *WFDC2* in Koreans leads to severe respiratory distress accompanied by bronchiectasis and rhinosinusitis": Supple figures and tables

### SUPPLEMENTARY FIGURES AND LEGENDS

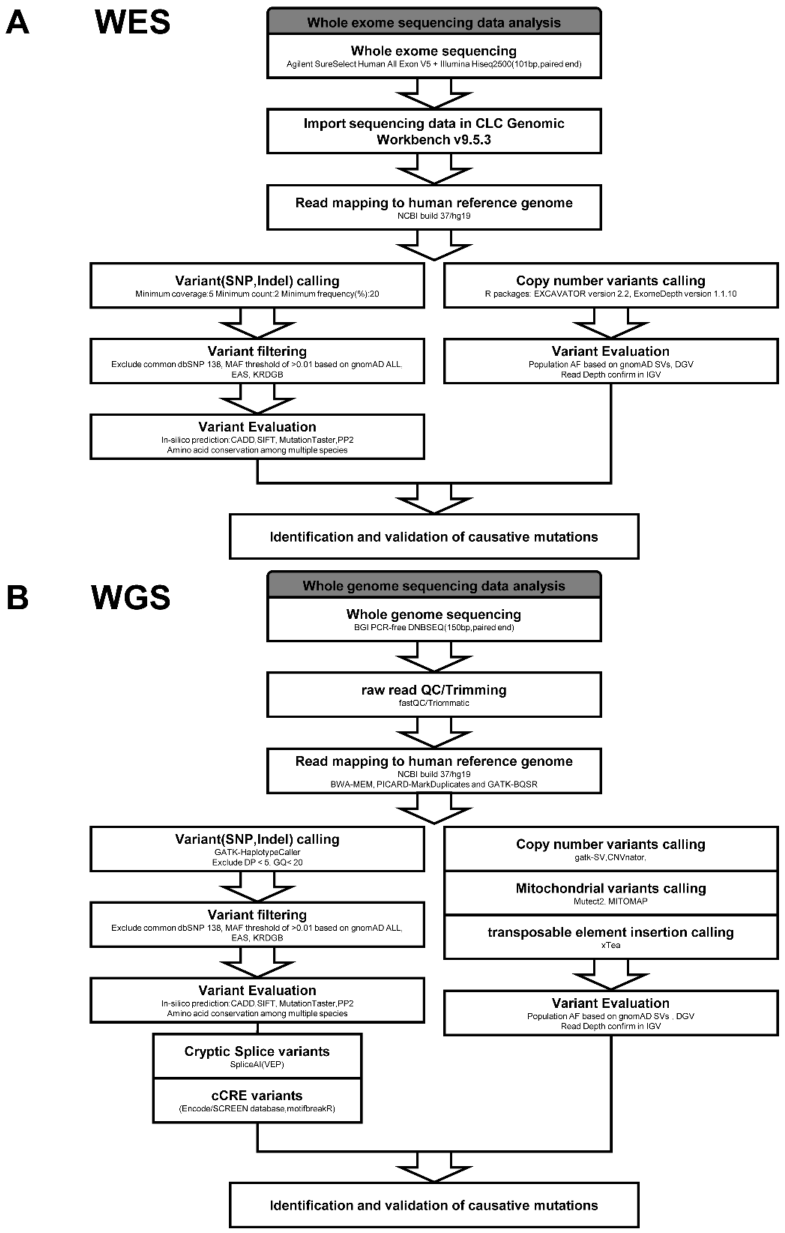


**Figure S1. Pipeline for sequencing data analysis aimed at novel gene discovery.**

(A) Data analysis pipeline for whole-exome sequencing. (B) Data analysis pipeline for whole-genome sequencing.

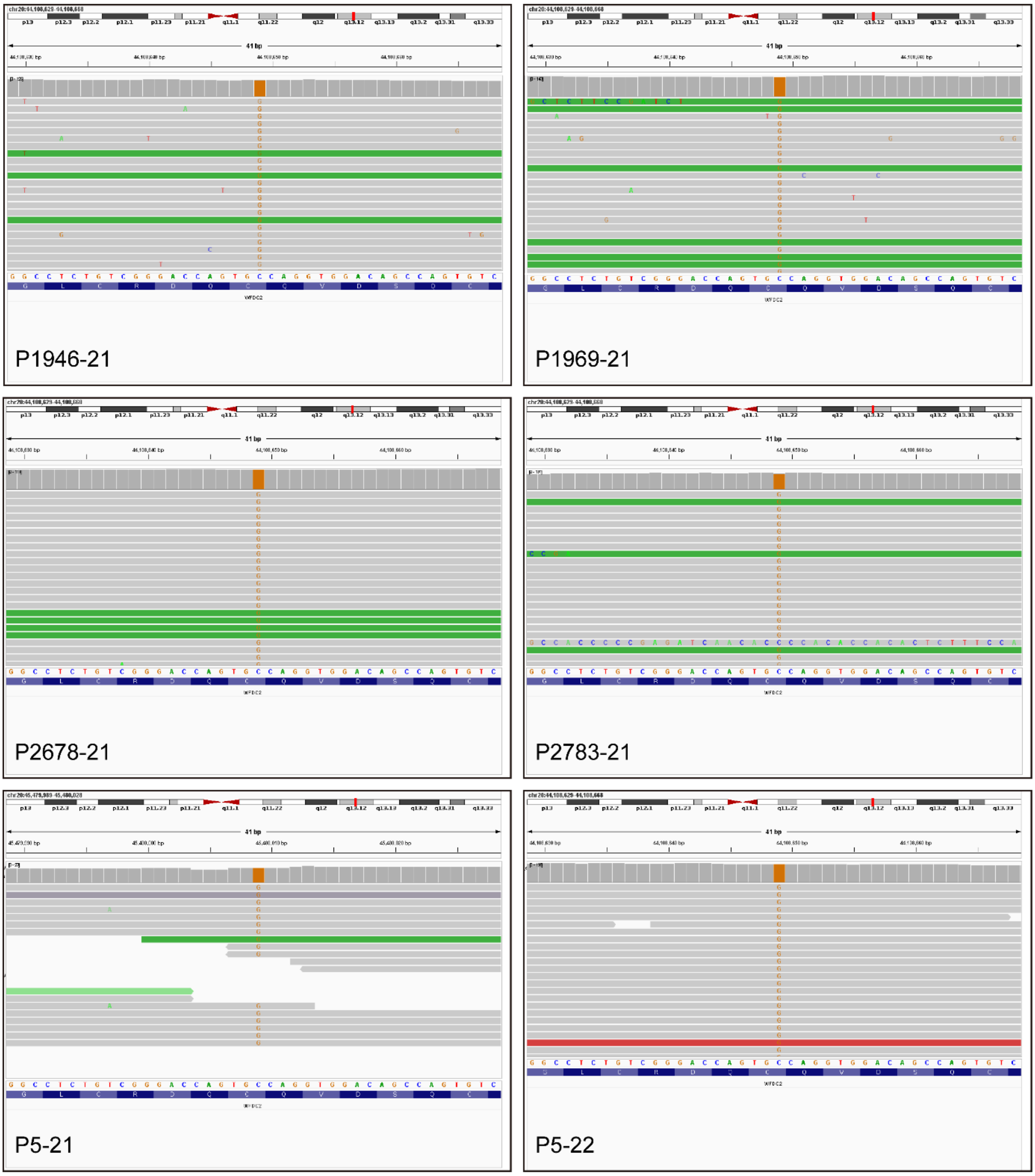


**Figure S2. Interactive Genomics Viewer (IGV) images of individuals with *WFDC2* c.291C>G, p.Cys97Trp variant.**

IGV images show the genomic region of individuals carrying the *WFDC2* c.291C>G, p.Cys97Trp variant, highlighting the mutation site and its surrounding sequence.

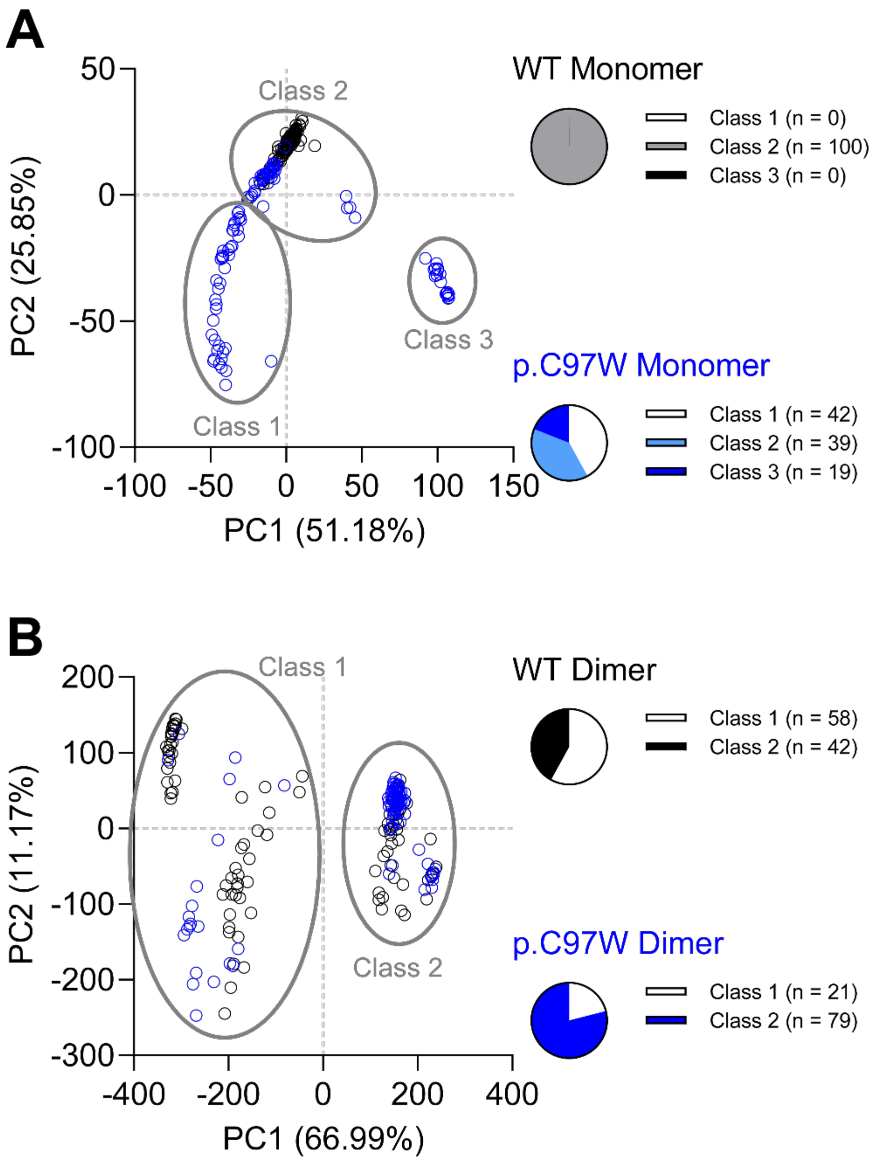


**Figure S3. Clustering analysis of the WFDC2 WT and p.C97W structures.**

(A) *K*-means clustering analysis of WFDC2 WT and p.C97W monomeric structures predicted using AlphaFold2[1] and ColabFold[2]. (B) *K*-means clustering analysis of the WFDC2 WT and p.C97W dimeric structures. Class 1 represents the *cis* conformation, and Class 2 represents the *trans* conformation.

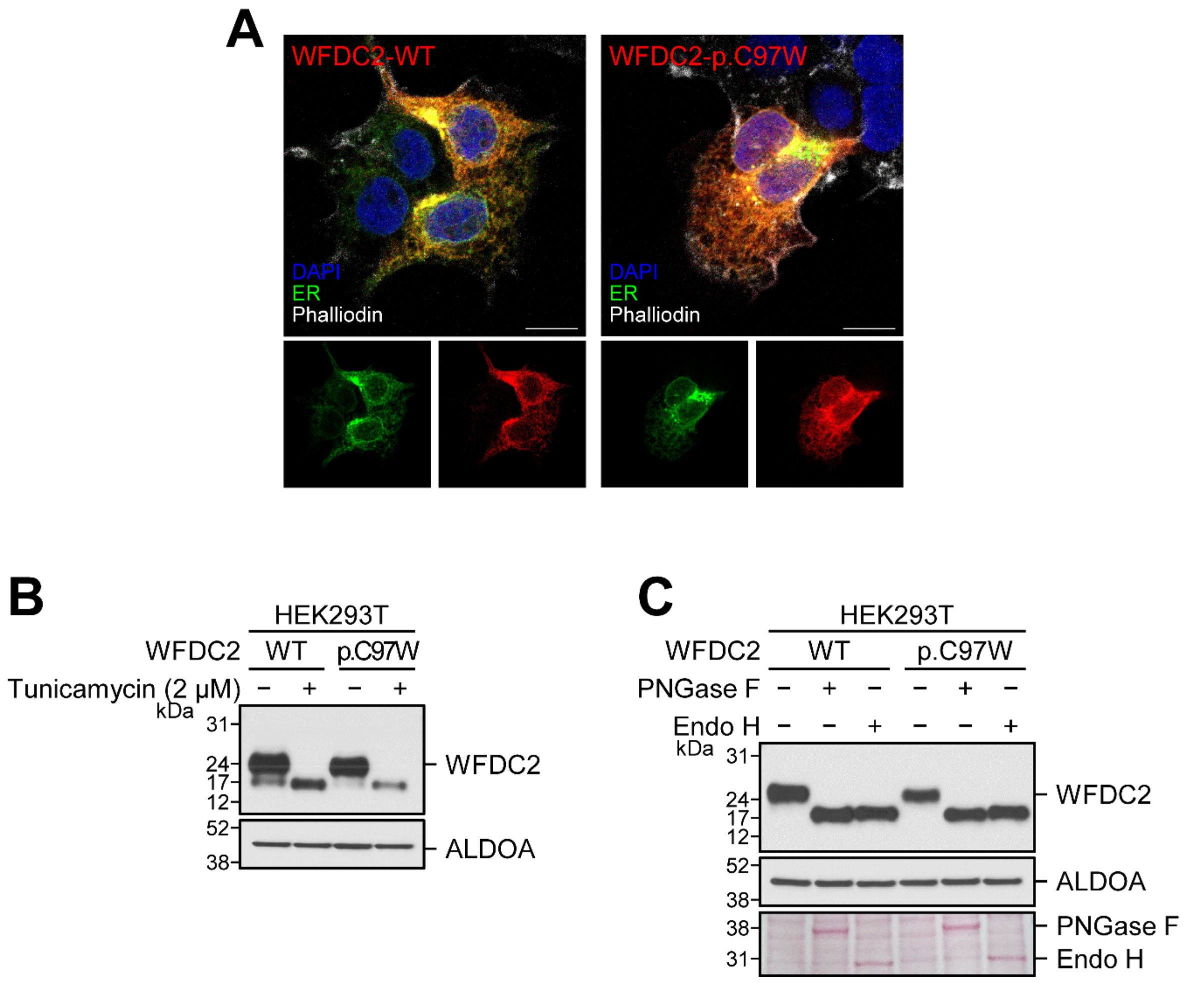


**Figure S4. Intracellular localization and glycosylation of WFDC2 WT and p.C97W.**

(A) Immunostaining of WT and p.C97W WFDC2 overexpression in HEK293T cells. pEYFP-ER was used as a reference gene. WFDC2 WT or p.C97W plasmid contained a 10xHIS tag in the C-terminus, which was used for staining WFDC2. pEYFP-ER was used as a reference marker for endoplasmic reticulum localization.

#### SUPPLEMENTARY TABLES

**Table S1. WFDC2 c.291C>G variant identified by whole exome/genome sequencing in five families**

| Nucleotide change[a] | Amino Acid Change | Zygosity | dbSNP[b] | gnomAD[c] | gnomAD EAS | KRGDB[d] | Mutation Taster[e] | PP2 Humvar[f] | SIFT[g] | CADD[h] |
|---|---|---|---|---|---|---|---|---|---|---|
| c.291C>G | p.Cys97Trp | HOM | rs780739822 | 0.00001611 | 0.0004051 | G:0.00181818 | D,D,D (0.999999,0.999999, 0.999978) | ProbablyDamaging (1) | Deleterious(0) | 23.6 |

[a]cDNA mutations were numbered according to the human cDNA reference sequence NM_006103.4(WFDC2). [b]dbSNP database(http://www.ncbi.nlm.nih.gov/SNP). [c]gnomAD (http://exac.broadinstitute.org/) software. [d]The Korean Reference Genome Database (http://coda.nih.go.kr/coda/KRGDB/index.jsp). [e]MutationTaster (http://www.mutationtaster.org/). A value close to 1 indicates a high “security” of the prediction [f]PolyPhen-2 prediction score HumVar ranges from 0 to 1.0; 0=benign, 1.0=probably damaging (http://genetics.bwh.harvard.edu/pph2/). [g]SIFT (http://sift.jcvi.org/) software. SIFT scores range from 0.0 (deleterious) to 1.0 (tolerated). [h]Phred-like scores (scaled C scores) on the Combined Annotation Dependent Depletion (http://cadd. gs. Washington. edu/home/).

**Table S2. Frequency of haplotypes consisting of eight SNPs in East Asians.**

| | rs537773418 (C>T) | rs2231617 (G>A) | rs1127497 (A>G) | rs80158178 (G>A) | rs2741566 (T>C) | rs707577 (T>C) | ↓ | rs76617043 (C>A) | rs192855884 (G>A) | | |
|---|---|---|---|---|---|---|---|---|---|---|---|
| **MAF** | 0 | 0 | 0.56 | 0.036 | 0.461 | 0.459 | | 0.179 | 0 | | |
| **Block** | | | | | | | | | | **Frequency** | **Count** |
| **H1** | C | G | **G** | G | **C** | **C** | | C | G | 0.3285 | 136 |
| **H2** | C | G | A | G | T | T | | C | G | 0.2729 | 113 |
| **H3** | C | G | **G** | G | T | T | | C | G | 0.1135 | 47 |
| **H4** | C | G | A | G | **C** | **C** | | C | G | 0.0797 | 33 |

The red arrowhead indicates the position of WFDC2 c.291C>G, p.Cys97Trp (hg19 chr20:44,108,649).

[a]JPT: Japanese in Tokyo, Japan

[b]CHB: Han Chinese in Beijing, China

**Table S3. LoF variants of *WFDC2* reported in gnomAD 4.1 (hg38).**

| Chromosome | rsIDs | Reference | Alternate | Protein Consequence | Transcript Consequence | Allele Count | Homozygote Count | Allele Number | Allele Frequency |
|---|---|---|---|---|---|---|---|---|---|
| 20:45469787 | | T | TGCTTGTCGCCTAG GCCC | p.Ala10ValfsTer3 | c.10_26dup | 2 | 0 | 1605282 | 1.25E-06 |
| 20:45469815 | | GC | G | p.Leu13SerfsTer12 | c.37del | 1 | 0 | 1610844 | 6.21E-07 |
| 20:45470387 | rs1470660005 | A | G | | c.80-2A>G | 3 | 0 | 1582216 | 1.9E-06 |
| 20:45470400 | | G | T | p.Glu31Ter | c.91G>T | 1 | 0 | 1588502 | 6.30E-07 |
| 20:45470406 | rs760216084 | A | ACTGG | p.Val35TrpfsTer9 | c.99_102dup | 2 | 0 | 1589656 | 1.26E-06 |
| 20:45470417 | | C | A | p.Cys36Ter | c.108C>A | 1 | 0 | 1590490 | 6.29E-07 |
| 20:45470427 | | C | T | p.Gln40Ter | c.118C>T | 1 | 0 | 1592640 | 6.28E-07 |
| 20:45470448 | | C | T | p.Gln47Ter | c.139C>T | 22 | 0 | 1595280 | 1.38E-05 |
| 20:45470474 | rs772005457 | C | A | p.Cys55Ter | c.165C>A | 3 | 0 | 1596508 | 1.88E-06 |
| 20:45470475 | | GC | G | p.Asp57ThrfsTer27 | c.168del | 3 | 0 | 1597264 | 1.88E-06 |
| 20:45470508 | | G | GC | p.Thr68HisfsTer8 | c.201dup | 1 | 0 | 1598946 | 6.25E-07 |
| 20:45470533 | rs371133876 | G | A | | c.223+1G>A | 6 | 0 | 1592602 | 3.77E-06 |
| 20:45479948 | | AG | A | p.Gly78ValfsTer6 | c.233del | 1 | 0 | 1614176 | 6.20E-07 |
| 20:45479958 | rs769309062 | C | A | p.Cys80Ter | c.240C>A | 2 | 0 | 1614240 | 1.24E-06 |
| 20:45479991 | | CCT | C | p.Cys93SerfsTer42 | c.276_277del | 1 | 0 | 1614254 | 6.19E-07 |
| 20:45480004 | rs1467482894 | C | T | p.Gln96Ter | c.286C>T | 14 | 0 | 1614234 | 8.67E-06 |
| 20:45480017 | | ACAGC | A | p.Gln102ValfsTer5 | c.303_306del | 1 | 0 | 1614194 | 6.20E-07 |
| 20:45480047 | | GC | G | p.Arg111AlafsTer22 | c.331del | 1 | 0 | 1614236 | 6.19E-07 |
| 20:45480084 | rs754010371 | CA | C | p.Asn123IlefsTer10 | c.368del | 7 | 0 | 1613972 | 4.34E-06 |
| 20:45480089 | | TC | T | p.Ter125GlufsTer8 | c.372del | 1 | 0 | 1613736 | 6.20E-07 |
| 20:45480095 | | G | T | | c.*1+1G>T | 1 | 0 | 1613542 | 6.20E-07 |
